# “I told myself, be bold and go and test”: Motivators and barriers to HIV testing among gays, bisexuals, and all other men who sex with men in Ghana – West Africa

**DOI:** 10.1101/2023.07.12.23292583

**Authors:** Gamji Rabiu Abu-Ba’are, Gloria Aidoo-Frimpong, Melissa Stockton, Edem Yaw Zigah, Samuel Amuah, Prince Amu-Adu, Richard Panix Amoh-Otoo, Laura Nyblade, Kwasi Torpey, LaRon E. Nelson

**Affiliations:** Behavioral, Sexual and Global Health Lab, School of Nursing, University of Rochester Medical Center, University of Rochester, Rochester New York; Department of Public Health Sciences, University of Rochester Medical Center, University of Rochester, Rochester New York; Yale AIDS Prevention Program (Y-APT), Center for Interdisciplinary Research on AIDS, School of Public Health/Medicine, Yale University, New Haven, CT; Department of Epidemiology, Gillings School of Global Public Health, University of North Carolina at Chapel Hill, Chapel Hill, NC; Behavioral, Sexual and Global Health Lab, West Africa Site, Jama’a Action, West Legon, Accra, Ghana; Priorities on Rights and Sexual Health, Accra, Ghana; Youth Alliance for Health and Human Rights, Kumasi, Ghana; RTI International, 701 13th Street NW., #750, Washington DC 20005, USA; Department of Population, Family & Reproductive Health, School of Public Health, University of Ghana, Legon-Accra, Ghana; School of Nursing, Yale University, New Haven CT, USA

## Abstract

Limited evidence from Ghana and SSA shows that despite consistently a disproportionately high burden of HIV, GBMSM infrequently often delay testing until the point of illness. We therefore used qualitative interviews to collect insights of experiences, motivators, and barriers to HIV testing among GBMSM. Two community-based organizations used snowball and convenience sampling to recruit 10 MSM for IDIs and 8 to 12 for FGDs. We transcribed, coded, identified and analyzed the relationship and commonalities between the participants’ responses. Under experiences with testing, 1) fear of HIV infection created a stressful HIV testing experience; and 2) friendly and supportive healthcare environment facilitated a positive experience in healthcare facilities. Motivators or facilitators of testing include: 1) the perception or belief that HIV testing is a HIV prevention strategy; 2) encouragement from friends and peers; 3) understanding risk associated with certain sexual behaviors such as transactional sex 4) education or information on HIV; 5) access to free testing and incentives; 6) early symptoms and provider recommendation. Barriers to HIV testing include: 1) negative community perceptions of HIV deter; 2) individual-level low risk perception or indifference about HIV infection; 3) location and cost; 4) inadequate testing availability; 5) Stigma at Healthcare facilities. The findings point to the need to address important issues around stigma, education, peer support and healthcare resources through interventions and research to improve HIV testing among GBMSM in the country.

## Introduction

Gays, bisexuals, and all other men who sex with men (GBMSM) are at heightened risk for HIV.[1, 2] As of 2021, GBMSM have 28 times the risk of acquiring HIV compared to heterosexual men.[3] In Ghana, 17.1% of GBMSM are estimated to be living with HIV; further, Ghanian GBMSM, along with other key populations, disproportionately account for new HIV infections.[4]. Ghana and other SSA, with the aid of international health organizations and programs have begun to focus on improving HIV outcome among key populations [1–5]. Yet several factors such as low HIV knowledge, condomless anal sex, discrimination and criminalization, and social isolation continue to hinder HIV prevention and care among GBMSM [1–10].

At the GBMSM level, limited studies in Ghana show that majority of GBMSM in the country have lower understanding of HIV prevention, and treatment processes [10–14]. For instance, a study among 137 GBMSM showed low mean scores for knowledge on STI and HIV transmission, prevention, and treatment [14]. As such, there is lower use of prevention tools such as condoms, lubricants and HIV testing, and in some cases, limited adherence to medication. For instance, condom use was higher when GBMSM had vaginal sex with women, (rel. f. = 0.75) than anal sex with men (rel. f. = 0.50) [14]. About 40% of did not use condoms during first sexual encounters, and even less (38%) continuously used condoms with sexual partners [14]. These increased risk behaviors and high HIV prevalence among GBMSM shows the need for increased HIV testing [10–16].

Regular HIV testing is key to both HIV prevention and linkage to care; HIV testing allows GBMSM to be aware of their HIV status, facilitating sexual-health decision making, access to vital prevention services such a pre-exposure prophylaxis (PrEP), as well as initiation of ART, improvement in HIV morbidity and mortality.[11, 12] While HIV testing practices are slowly increasing among GBMSM in Africa,[2, 19] there is a need to understand the factors the affect HIV testing behaviors for this vulnerable group, and to begin to address such factors [15,16].

In addition to low HIV knowledge, and access to HIV prevention and care services, high HIV and sexual stigma and discrimination affect HIV testing among GBMSM [15,16]. Recognizing the centrality of stigma in HIV testing practices, this study employs the Health Stigma and Discrimination Framework (HSDF) to unpack GBMSM’s experiences across the socio-ecological model.[20] HSDF posits that the stigmatization process unfolds across the socio-ecological spectrum in the context of a health condition, such as HIV care, and ultimately impacting uptake of testing, adherence and treatment, incidence, morbidity and mortality among affected groups [20]. Consistent with the HSDF, Ghanaians in general have high misconception of HIV and treatment, and view HIV like a death sentence, hence, many GBMSM have fear of a positive diagnosis.[4, 9–16]. Further, same-sex sexual behaviors can be treated as criminal in Ghana and a recent national anti-LBGTQ political rhetoric hamper both service delivery and engagement in HIV prevention programming [21, 22]. As such, GBMSM face stigma at the community level and health care facility levels, which can manifest in the form of gossip, verbal and physical abuse, blame, shame and ultimately poor-quality of healthcare services.[9, 10, 23]. All these manifestations of stigma occur at higher levels for GBMSM who also have gender non-conforming expressions (example, men who act it in ways that will be considered feminine in the Ghanaian context), not only from community but healthcare providers, but at the interpersonal level between GBMSM peers [9, 10, 23].

The internalization of stigma associated with HIV, sexual status, and gender expressions, can combine with limited HIV knowledge and other factors to reduce interest in HIV testing, prevention and care services among GBMSM. Yet, there is limited research on the HIV prevention experiences of GBMSM or into the factors that motivate or hamper HIV testing behaviors among GBMSM in Ghana. A better understanding of these factors may facilitate access to comprehensive HIV prevention services and ultimately reduce disparities in HIV testing and incidence.[2] This qualitative study therefore, uses the lenses of the HSDF to understand HIV testing experiences of Ghanaian GBMSM, and factors that motivate or block willingness to test for HIV.

## Methodology

### Design

This study constitutes a part of a formative phase for a randomized controlled trial (RCT) that aims to assess the feasibility, acceptability, and potential effect size of a multi-level intervention to reduce intersectional HIV, gender nonconformity, and sexual stigma and to increase HIV testing among GBMSM in Ghana.[12] Here, we provide a summary report of the study approach, a detailed description has been published elsewhere [10–12] The assessment phase reported in this paper used a multi-method phenomenological approach, which combined focus group discussions (FGD) and in-depth interviews (IDI) to understand experiences of HIV testing among GBMSM in Ghana. The FGDs included questions pertaining to HIV testing experiences, motivators and facilitators to HIV testing, barriers to HIV testing at the intra and interpersonal levels, community levels, and healthcare facility level. In addition to the FGD questions, the IDI also inquired about positive and negative experiences around HIV testing and diagnoses.

### Sampling and Data Collection

Two GBMSM-led community partners in Accra and Kumasi employed a combination of convenience and snowball sampling to recruit GBMSM in Ghana. FGD participants were 18 years or older, male at birth, self-identified as men, and had sex with other men within the previous six months. In addition, in-depth interview (IDI) participants also self-disclosed that they were living with HIV. The interviewers were individuals from the partner organizations who self-identified as gay and bisexual men from the respective cities and had received a data collection and human subject research training. They invited GBMSM to participate in the IDIs and FGDs at their respective organizations. Eight FGDs and 10 IDIs were conducted. Both the IDIs and FGDs were primarily conducted in English, although two FGDs were conducted in Twi. The interviewers digitally recorded and transcribed all IDIs and FGDs. The research team reviewed both English and Twi audio files to ensure consistency. The research team reviewed the first several IDIs, provided the interviewers’ feedback, and revised the interview guides as necessary.

### Ethical Approval

The institutional review boards of Yale University, Noguchi Medical Research Institute, the Ghana Health Service, and the University of Toronto approved the study. All study participants signed an informed consent form and received a token for their participation. Only community level partners had direct access to participants, they only provided aggregated data without any information for coding and analysis. Hence, no information harmful to participants is accessible to any co-authors in this manuscript.

### Data analysis

Seven members from the research team and partner organizations coded the data line by line. First, all members reviewed the transcripts to identify relevant concepts which informed the construction of a code book. Using the code book, two coders independently coded the same transcripts and compared their codes to reach a consensus in terms of discrepancies in a meeting. We then combined all codes into broad classifications of participants’ experiences and perspectives. The research team examined the resulting groups of codes across all transcripts and reviewed to confirm their comprehensiveness and representation.

## Results

We identified and analyzed the relationship and commonalities between the participants’ responses to understand HIV testing experiences of GBMSM, and factors that motivate or block willingness to test for HIV. Under experiences with testing, 1) fear of HIV infection created a stressful HIV testing experience; and 2) friendly and supportive healthcare environment facilitated a positive experience in healthcare facilities. The following factors served as motivators for or facilitated testing: 1) the perception or belief that HIV testing is a HIV prevention strategy; 2) encouragement from friends and peers; 3) understanding risk associated with certain sexual behaviors such as transactional sex 4) education or information on HIV; 5) access to free testing and incentives; 6) early symptoms and provider recommendation. We also identified the following barriers to HIV testing: 1) negative community perceptions of HIV deters; 2) individual-level low risk perception or indifference about HIV infection; 3) location and cost; 4) inadequate testing availability; 5) Stigma at Healthcare facilities.

### Experiences of HIV Testing

1. Fear of HIV infection created a stressful HIV testing experience. Many FGDs participants described their experiences of the testing process and their feelings while getting tested. A few participants described the process as quite “traumatic”, and scary as they were unsure about what the test results will be. One participant reflected on his experience, “I could remember when I went to do my HIV test; it was very scary; I was like “wow ooh my gosh! What if I am HIV positive? What will happen next?” So when I did my test the results didn’t even come, but I was crying; I was crying and I was so scared because I was like “Eish! so if I am HIV positive would I die? I would die because I can’t take medicine for the rest of my life” A whole lot was on my mind. Even within that five to ten minute, I was sweating because I didn’t know what the results would turn out to be, so I was like “wow Oh God help me, don’t let it be that I am HIV positive” So you know it was like a whole lot.” (FGD 1) The rest of his group echoed this sentiment. Another participant shared, “Being HIV positive is not something you should be happy with, so when I was going for the test I was having this panic attacks and anxiety” (IDI3)
2. Friendly and supportive healthcare environment fwacilitated a positive experience in healthcare facilities Participants highlighted the positive experiences associated with their testing experiences in both public and private facilities. Privacy and free testing were identified as central to their positive experiences: “there wasn’t an obstacle because HIV test is free.” (FGD 1). Others shared that the process of testing was quite easy, with nurses being friendly and “treating them equally.” Conversations with healthcare providers served as a source of encouragement and comfort to participants, helping them to realize and understand their risk of an infection. One participant reflected: “Because I was having these few friends and they sometimes do meetings and they sometimes bring nurses and we talk about HIV and stuff and they realize . . . they [Nurses] made us know that we are really at risk. So, it prompted me to give it a chance to do the test”

### Motivators and Facilitators of HIV testing

Participants who had tested for HIV discussed their reasons for getting an HIV test. Participants highlighted peer influence, their sexual orientation, sexual behaviors, receiving information about HIV, work requirements, and having symptoms as the significant reasons for testing.

1. Perception of HIV testing as a prevention strategy served as a motivator for testing. Most of the participants had tested for HIV, with a few participants revealing that their most recent test was a week prior to the FGD. There were differing opinions related to HIV testing for prevention. Some participants were convinced that based on their lifestyle as GBMSM, they were more at risk of contracting HIV; thus, it was imperative that they know their HIV status: “As a sexually active person and an MSM, am at a very higher risk… So anytime I get the chance I want to test.” (FGD 1) Participants shared that testing provided peace of mind and helped protect themselves and others within their networks from being infected. One participant mentioned: “I think the very best thing that could happen to someone who is MSM is to test for HIV like in very simple and short words. That’s what I tell people because once you know yourself and you know the risk you get exposed to once a while or you know your identity, the best thing for you is to test for HIV.” (FGD3)
2. Encouragement from friends and peers serve as a motivator for testing. Participants had overall positive experiences of peer support. Getting tested resulted from the encouragement and support from their peers. This support included providing information about HIV, sharing their own testing experiences, accompanying them to get tested at the healthcare facility. One participant shared, “A friend of mine who had gone in to test his HIV status encouraged me to also test and know my status.” (FGD2). Another participant explained: “I have friends who are peer educators and they kept telling me that I should come so in the world AIDS day two years ago I tested” (FGD1)
3. Understanding risk associated with sexual behaviors such as transactional sex served as a motivator for testing. Testing for HIV, according to a few participants, was a critical need for GBMSM. Participants believed that they had a greater biological risk of HIV acquisition because of anal sex, thus being or identifying as GBMSM was enough of a reason to get tested: “Once you have accepted you are an MSM, you need to check your status. I’m saying this because if heterosexuals are getting tested to know their status so why not you the MSM. I believe as MSM we should get tested.” (FGD 3). Another participant also described the perceived risk of being GBMSM and the need to get tested: “So as an MSM person living, it’s always good for you to at least go for your test every three months or every six months because you yourself know that you are doing something that puts you at risk. So, your life is always at risk.” (IDI 4) Similarly, other men acknowledged this sentiment and highlighted that other sexual behaviors of GBMSM such concurrent partnerships put them at risk, thus necessitating consistent HIV testing. One participant shared his thoughts: “I don’t have one sexual partner. I have multiple sexual partners and as a guy or as an GBMSM having multiple sexual partners, I think it’s better for me to test to know my status.” (FGD3). Other participants also revealed that a preference for unprotected sex served as a motivating factor to get tested: “What helps me to always get tested is because I always know that I am an GBMSM and sometimes I do love to have unprotected sex, so I always like to go for testing at least every three months.” (FGD2) Transactional sex, which serve as a source of income for some participants, was discussed as a reason to get tested. Participants who engaged in transactional sex described HIV testing as a way of protecting themselves and their clients. A participant explained why being involved in sex work required that they get tested often: “I got tested because of my job (sex work) Yes, I see it as job because I and some friends that I know depends on this to survive. Many of us don’t work so exchanging sex for money is what keeps us moving.” (FGD 1). Another participant expressed his support by saying “I know the kind of job I do and I have no plans of stopping anytime soon. So, I think frequent testing is key” (FGD1) Perceived partner unfaithfulness also motivated HIV testing. Few participants highlighted the lack of trust in their partners as a factor that motivated them to test regularly: “I have a partner and I don’t trust him for being faithful, so testing is important to me as an GBMSM.” (FGD 3)
4. Education/Information on HIV motivated participants to test. Receiving information on HIV through peer navigators, clinics, and health fairs was critical to HIV testing among the participants. Many of the FGD participants shared that they ultimately decided to test after receiving simple, but detailed information about HIV transmission and risk. “At first, I didn’t want to test, but I changed my mind when I had education from a peer educator. That’s how I had my first test.” (FGD 1).
5. Access to free testing and incentives facilitated testing Monetary incentives added to free testing opportunities also viewed was a critical motivator for testing: “I think it was a research program or something like that and there was money involved, I think they were giving ₵30.00 or something and I wanted to get the money and I got there and I was tested” (FGD 1). Another participant shared “I got tested because I wanted the 40 cedis trust me, it’s the truth I’m telling you” (FGD2)
6. Early symptoms and provider recommendations facilitated testing Receiving a recommendation or instructions from a healthcare provider to test for HIV was critical to HIV testing decision-making. A few individuals described deciding to test per their doctor’s recommendation: “The doctor said I should go and do it so I just decide to do it.” (FGD 2). For others, the recommendation was related to an undiagnosed disease, with the doctors suggesting an HIV test as a last resort. A participant shared: “So there was a time the doctors were not figuring what was the sickness. So one of the doctors told me to go and run an HIV test. So I went [to get tested].” FGD 4

Experiencing symptoms such as fevers, general body weakness, weight loss, loss of appetite, headaches, rashes on the body, and discharges from the penis prompted some of the men to get tested: “Sometimes I take my bath and my body itches me; That alone was enough for me, and I felt like I was also losing weight a bit. Though I am not the fat type, but I felt I lost weight a little bit. Somebody would sometimes see you and would act like “are you sick?” and that kind of stuff so it alerted me and it made me feel like okay let me go and test and see why I am having this kind of feeling. That’s the reason why.” (FGD 2). Another participant also explained why they got tested: “And you know once in a while you feel some rashes and you feel a bit sick and that kind of stuff and sometimes you feel like you have lost appetite and those kinds of things. So I really wanted to test so that I would really see what is really wrong with me.” (IDI2) Another participant also shared the symptoms they had which encouraged them to go get tested: “I had a discharge from my penis and headache” (IDI3).

A few participants shared that they got tested due to work policies or as an entry requirement for certain occupations such as the military and nursing school; as one participant mentioned: “Because of the nature of my work, I have to get tested. I am a caterer.” (FGD 2). Another participant described their experience with these mandatory testing policies: “Well it’s the school’s policy, so you have to do it whether you like it or not. Before you go to nursing school you have to go for a medical report, just for you to be fit for the training school. So I got tested last two years. I was like, this thing would come out and what if I’m positive? Because I had never tested myself before, so I was thinking a lot, but it came out that I was not reacting. I was like “Hurray!”” (FGD 4) Barrier to HIV testing

1. Negative Community perceptions of HIV deters testing Participants discussed how community understanding of HIV is critical to prevention needs. HIV as a disease was usually associated with immoral behaviors, as a punishment for deviant behaviors related to sex or contracted by only certain groups such as MSM or commercial sex workers. Participants discussed that these prevalent community beliefs about HIV led to stigma, which they revealed impacted prevention activities such as testing. One participant reflected: “They say it’s [HIV] a satanic disease; you got this disease from fornication and whoring, and all sorts of stuff. On my first day I told myself “Yaw! Be bold and go and test, if you are negative then you are negative, and if you are positive, you know what to do.” (FGD2) Another participant described, “In my community, they talk bad about HIV especially when you are gay. They say it is MSM or gays who usually get infected with this kind of disease. So they talk bad about that HIV.” FGD 4
2. Individual level low risk perception / indifference about HIV infection hinders testing In a world of competing health risks, some participants did not perceive HIV to carry a severe health risk. A few FGD participants believed that many other diseases could kill them, thus, there was no need for the extra focus on HIV. One participant explained, “Let me say, I am not interested in testing or I have never tested or nothing because as I said I work in the health sector and my philosophy in life is that, all die be die cos working in the health sector and looking at the things that are killing people in Ghana, HIV is not top 10 killer disease”. The most killer disease is RTA, Road Traffic Accidents or malaria those days so I don’t see myself bothering about HIV or whatever like to me I am in the hospital, I don’t, if HIV will kill you, it will kill you for even recently corona [COVID-19] is in and its killing people so what’s the big deal, you understand it. HIV is still there so I don’t, I have never tested and I don’t think I will ever test.” (FGD3)
3. Distance and transportation cost as barriers to HIV testing The cost of transportation to the centers to get tested served as a barrier to getting tested. Participants discussed that since testing was free, the challenge usually involved paying for travel to the health centers. One participant explained: “I will say transportation will be the only cost but there were no charges at the facility. And it was free. All I need to do is to be at the facility and the test will be done. So, the only cost will be moving from my home to the facility to get tested.” (FGD 3).
4. Inadequate testing availability Shortage and inadequate testing kits were highlighted as barriers to testing by some participants. Participants mentioned that despite their readiness to get tested, there was a lack of testing kits available at the facilities they visited over multiple months. A participant revealed: “It was the shortage of test kits …I went to the facility and I was told there wasn’t any test kit.” (FGD 2).
5. Healthcare level sexual stigma deters GBMSM from testing for HIV Some FGD participants discussed the lack of MSM-friendly nurses and stigma in the form of healthcare providers’ demeaning comments towards MSM because of their engagement in same-gender partnerships and sexual activities. A participant described their experience: “I think there was this one facility that my partner and I visited at that time. I heard some comments from one of the nurses that made me felt uncomfortable so we refused to get tested at that time so we had to leave… One of the nurses said your people are here so the friendly nurse responded I know. So, she added how can a nice guy like this be Gay?” (FGD 2). Another person recounted; “the health provider told him straight away, do you do men? All of a sudden he was thrown aback, why you ask such a question because it is not only MSM who contract HIV. So he tried to question the health provider and the health provider said that immediately I saw you I knew you were one of them”. They come here all the time and we test them, I know how they behave.” FGD4Others added they do not to go to facilities with no friendly providers; “I know a lot of HIV testing sites that are not MSM friendly because they were not having friendly Nurses. I had to move from my place to another place for the HIV test.” IDI 5“

## Discussion

Limited evidence from Ghana and SSA shows that despite consistently a disproportionately high burden of HIV, GBMSM infrequently test for HIV and often delay testing until the point of illness [22–26]. This study therefore provides qualitative insights of experiences, motivators, and barriers to HIV testing among GBMSM. The findings point to important issues around stigma, education, peer support and healthcare resources that affect HIV testing and can inform intervention to increase testing among GBMSM in the country.

We established that stigma associated with HIV, and same-sex intercourse increases negative experiences with testing and dissuade GBMSM from testing. Consistent with the Health Stigma and Discrimination Framework (HSDF), accounts of participants show how intersectional stigma at the healthcare-, community-, and individual-levels affects testing experiences and discourage GBMSM from testing [20, 27]. At the community-level, GBMSM recount the association of HIV with sin, same-sex intercourse and sex work; at the healthcare level they experience negative comments such as gossips, and insults around sexual behavior. The accounts by participants corroborates the limited findings in Ghana and West Africa that show community level HIV and same-sex intercourse stigma as factors that affect HIV testing among GBMSM in the region [10, 23, 28]. As explained by the HSDF and previous findings, participants in this study internalized the stigma and begin to develop fear for HIV, thus experiencing stress during HIV testing, and or avoid testing in its totality [10, 23, 28].

Despite the negative effects of stigma, we found that positive healthcare environment can provide better experiences and encourages HIV testing among GBMSM. Participants who reported positive experience with testing mentioned the professionalism and confidentiality of providers at the testing sites as factors that contributed to their positive experience. Likewise, those who avoid testing sites mentioned that stigma from providers prevent them from testing. Our findings on positive healthcare environment as a facilitator for testing confirms findings among GBMSM elsewhere [29,30]. It also explains reported GBMSM preference for friendly and GBMSM led healthcare facilities, compared to government facilities where stigma appear more rampant [32, 32]. We therefore reaffirm the importance of implementing interventions that seek to alter stigma and negative experiences of GBMSM in public healthcare facilities, to increase HIV testing [12, 13, 23].

Our findings also point to the importance of increasing education and proper information channeling to GBMSM in Ghana to increase HIV testing. The participants who expressed understanding of HIV risk associated with same-sex intercourse, HIV transmission, and the role of HIV testing mentioned those as factors that informed their decision to test for HIV. Conversely, participants who expressed a lack of awareness, individual level low risk perception and or indifference about HIV infection did not test for HIV. Whereas limited interventions or studies focused on GBMSM in Ghana and sub-Saharan African countries, we previously implemented an adapted Many Men Many Voices intervention in Ghana that addressed knowledge and capacity around HIV risk, transmission, prevention and care among GBMSM [13, 15, 33]. The findings of this study support our initial evidence from the Many Men Many Voices intervention that showed that HIV testing and readiness to test for HIV increased after receiving education and information around HIV [13, 15, 33]. Similar findings appeared in some other studies in different parts of West African and SSA regions [34, 35].

Moreover, per our findings, peers can serve as promoters of HIV testing among GBMSM. As evidenced in this study, some participants tested for HIV because they were referred to testing by their friends and acquaintances, as well as peer educators who reach out to GBMSM to encourage testing. These findings confirm studies elsewhere where by a peer support intervention implemented among GBMSM contributed to address individual level challenges and increasing HIV testing [35,37].

Additionally, whereas there have been recent efforts through several programs to increase HIV testing among GBMSM in Ghana and SSA countries, access still remains affected by cost and location of facilities for some GBMSM [3, 4]. Some participants indicated that they could not afford to travel to the location where they can access HIV testing, others said they could not pay for HIV testing nearby. Therefore, emphasizing the urgent need to continue to increase support for programs that provide targeted HIV testing services to GBMSM free of charge. The issue of location and travelling can also be remedied by using newly available HIV self-testing which was recently introduced in Ghana [15, 38]. HIV self-testing, if implemented through existing peer networks, can provide ease access to GBMSM irrespective of their locations [15].

We implore researchers to consider the following limitations in interpreting our findings. As typical of nonprobability sampling, the convenience and snowball sampling used in the data collection limits its generatability for GBMSM in Ghana [39]. We also recruited GBMSM specific to Accra and Kumasi, the two largest urban settings in Ghana, thus findings may not represent other localities in Ghana. Additionally, this study was conducted to inform the development of an intervention to address intersectional stigma and increase HIV testing, thus limiting the scope to testing and no other aspects of care [12]. Hence, the findings should be used in addition to others findings on HIV testing among GBMSM in the geographical region. Nonetheless, the study provides an updated important insight to barriers, motivators and facilitators of HIV testing, which when considered will help in understanding ways to increase access to HIV testing among GBMSM in Ghana and ultimately helping Ghana reach its 95 95 95 HIV targets.

## Data Availability

The datasets used and/or analyzed during the current study are not publicly available due to our ethical and legal requirements related to protecting participant privacy and current ethical institutional approvals but are available from the corresponding author on reasonable request pending ethical approval.

